# Modifiable lifestyle activities affect cognition in cognitively healthy middle-aged individuals at risk for late-life Alzheimer’s Disease

**DOI:** 10.1101/2022.03.14.22272340

**Authors:** Amy Heneghan, Feng Deng, Katie Wells, Karen Ritchie, Graciela Muniz-Terrera, Craig W Ritchie, Brian Lawlor, Lorina Naci

**Author notes:** These authors contributed equally to this work (Amy Heneghan, Feng Deng). **Corresponding author:** Lorina Naci, School of Psychology, Trinity College Institute of Neuroscience, Global Brain Health Institute, Trinity College Dublin, Dublin, Ireland.

## Abstract

It is now acknowledged that Alzheimer’s Disease (AD) processes are present decades before the onset of clinical symptoms, but it remains unknown whether lifestyle factors can protect against these early AD processes in mid-life. We asked whether modifiable lifestyle activities impact cognition in middle-aged individuals who are cognitively healthy, but at risk for late life AD. Participants (40–59 years) completed cognitive and clinical assessments at baseline (N = 206) and two years follow-up (N = 174). Mid-life activities were measured with the Lifetime of Experiences Questionnaire. We assessed the impact of lifestyle activities, known risk factors for sporadic late-onset AD (Apolipoprotein E _□_4 allele status, family history of dementia, and the Cardiovascular Risk Factors Aging and Dementia score), and their interactions on cognition. More frequent engagement in physically, socially and intellectually stimulating activities was associated with better cognition (verbal, spatial and relational memory), at baseline and follow-up. Critically, more frequent engagement in these activities was associated with stronger cognition (verbal and visuospatial functions, and conjunctive short-term memory binding) in individuals with family history of dementia. Impaired visuospatial function is one of the earliest cognitive deficits in AD and has previously associated with increased AD risk in this cohort. Additionally, conjunctive memory functions have been found impaired in the pre-symptomatic stages of AD. These findings suggest that modifiable lifestyle activities offset cognitive decrements due to AD risk in mid-life and support the targeting of modifiable lifestyle activities for the prevention of Alzheimer’s Disease.

## Introduction

Dementia is a growing pandemic that presents profound challenges to health care systems, families, and societies throughout the world. Alzheimer’s Disease (AD), the most common etiology of dementia, is characterized by relentless neurodegeneration and accelerated cognitive decline in the years following presentation of clinical symptoms. It is now accepted that AD pathological processes are present decades before clinically relevant symptoms appear [1-2]. In the absence of effective pharmacological treatments, there is an immediate need for early identification, intervention and risk burden modification [3-4] for AD.

Growing evidence suggests that up to 40% of all dementia cases are associated with known lifestyle-related modifiable risk factors, such as alcohol consumption, obesity and hypertension among others [4]. As exposure to most of these risk factors begins decades before dementia onset, interventions must be implemented in mid-life [5-7]. Mid-life, thus, presents a critical and unique window for disease-altering interventions, before the manifestation of substantial brain damage. However, the indicators of AD in mid-life, and the impact of modifiable lifestyle factors on the incipient disease process remain poorly understood.

AD is neuropathologically characterized by the accumulation of beta-amyloid (Aβ) and hyperphosphorylated TAU (pTau) [8-9]. The etiology of the sporadic form of AD remains poorly understood. However, recent studies suggest that tau deposition, is a key etiological factor that presages sporadic AD [10-12]. Analyses of thousands human brains across the lifespan show that tau pathology begins about a decade before formation of Aβ plaques [13]. Additionally, tau pathology, but not Aβ, correlates with progressive grey matter loss [14] and cognitive impairment [15].Neuropathological findings show that tau pathology starts in late young adulthood and early midlife [30–40years], in subcortical nucleus locus coeruleus (LC) [9] —the key brain site for the production of noradrenaline— then spreads initially to the transentorhinal and entorhinal cortex [8], to the hippocampal and neocortical association cortex, and finally throughout neocortex. By contrast, Aβ plaques are usually first deposited in the association neocortex of the temporal lobe, and then extend throughout the cortex, including subcortical structures, with disease progression [13]. Therefore, brain changes related to sporadic AD in early midlife (i.e. 40-50 years) are likely associated with neurodegeneration from to pTau deposition and, in later age groups, Aβ plaques additionally contribute strongly to underlying neurodegeneration.

Studies on preclinical AD have used risk stratification approaches to investigate early, preclinical changes. Key risk factors include the Apolipoprotein E (APOE) _□_4 genotype [16-17], the main genetic risk factor for sporadic late onset AD in the Indo-European population [18], and family history (FH) of dementia [16, 19-20]. Several dementia risk scores incorporating lifestyle risk factors have been devised [21-23]. Amongst them, the Cardiovascular Risk Factors Aging and Dementia (CAIDE) score has been optimized for middle-aged populations [23] and has been validated in a large US population followed longitudinally over 40 years [24].

The PREVENT Dementia Program [25] initiated in 2013, is a prospective study of the cognitively healthy middle-aged children of persons with dementia, designed to seek out clinical and biological changes, which may subsequently be used as short-term outcome measures for midlife secondary preventions. Studies of this and other similar cohorts of cognitively healthy midlife individuals have related these three risk factors to a range of structural and functional brain changes, including APOE _□_4 genotype to loss of volume in the hippocampal molecular layer [26], cerebral hyperperfusion [27-28], reduced grey matter volume in the right hippocampus, precentral gyrus, and cerebellar cortex [29], and to decreased cortical thickness in the frontal cortex [30]; FH to volumetric alterations in hippocampal subfields and to disrupted white matter integrity [27, 31]; and, CAIDE to whole brain atrophy [32-34] and to hippocampal volume loss [31, 34]. All three risk groups have also been found to impact cognition in mid-life. APOE _□_4 genotype has been significantly associated with improvements in verbal, spatial and relational memory [35], immediate recall [36] and form perception [34], FH with poorer verbal processing and memory performance in participants under 65 years [37], and poorer executive function [38], and higher CAIDE scores with impaired verbal and visuospatial functions, and short-term (conjunctive) memory [34-35], and impaired executive function [39]. Taken together, these studies have established that risk for late-life AD, including risk that incorporates lifestyle factors, has a significant impact on the brain health and cognition of middle-aged individuals who are presently cognitively healthy.

As a multidimensional construct, lifestyle has a multipronged impact on cognition and the brain. By contrast to aforementioned cardiovascular risk factors captured by the CAIDE score, several lifestyle activities have been found to protect brain health and cognition in later life. Factors, such as education and occupational complexity have been associated with preservation of cognitive function in order adults [40] and reduced symptom severity in Alzheimer’s disease [4, 41-43], a phenomenon known as “cognitive resilience”. These factors are thought to explain why, in late-life Alzheimer’s disease, the level of cognitive impairment shows substantial variability even when accounting for key pathologies including Aβ and pathological tau [1, 44]. One prominent account of the biological mechanisms mediating the relationship between stimulating lifestyle activities and cognitive resilience in AD [45-46] suggests that environmental enrichment upregulates the noradrenergic system which originated in the locus coeruleus, that otherwise depletes with age [47] and AD pathology [10], leading to compensatory brain mechanisms for cognitive function.

While epidemiological evidence strongly suggests that education and occupation contribute to cognitive resilience [48], there is a renewed interest in the additional contribution of other activities undertaken in mid-life, given their potential modifiability. For example, Gow et al. (2017) [49] and Chan et al. (2018) [40] used the Lifetime of Experiences Questionnaire (LEQ) and found that physically, socially and intellectually stimulating lifestyle activities undertaken in mid-life, independently of education, help to maintain late-life cognitive performance in older adults, after adjusting for childhood cognitive ability [49]. However, it is not known whether the protection conferred by lifestyle activities, other than education, offsets the impact of AD risk in mid-life, or whether it builds up gradually over time with its benefits viable only in late life or relative to established AD pathology. The answer to this question is critical for identifying interventions that target modifiable factors for the prevention of Alzheimer’s disease from the earliest life stages.

To address this gap, we investigated the impact of interactions between three risk factors for late-life AD (APOE _□_4 genotype, FH and CAIDE) with lifestyle activities on cognition, independently of sex, age and years of education. These relationships were investigated in a large cohort of cognitively healthy middle-aged individuals, assessed at baseline (N=206), and at two years follow-up (N=174). To relate our findings to previous studies, we used the LEQ [50], the same instrument as in aforementioned studies, to evaluate lifestyle activities specific to mid-life, yielding two composite factors, (a) occupation and managerial responsibility, and (b) physical, social and intellectual activities. Our hypothesis was that there would be a significant association between mid-life lifestyle factors and cognitive performance in domains already shown in this cohort to be affected by risk for late-life AD.

## Methods

### Participants

PREVENT is an ongoing longitudinal multi-site research programme based across the UK and Ireland, seeking to identify early biomarkers of AD and elaborate on risk-mechanism interactions for neurodegenerative diseases decades before the cardinal symptoms of dementia emerge. Its protocol has been described in detail elsewhere [25]. In the first PREVENT programme phase, participants were recruited at a single site, via the dementia register database held at the West London National Health Service (NHS) Trust, of the UK National Health Service, the Join Dementia Research website (https://www.joindementiaresearch.nihr.ac.uk/), through public presentations, social media and word of mouth. Procedures involving experiments on human subjects were done in accord with the ethical standards of the Institutional Review Board of Imperial College London and in accord with the Helsinki Declaration of 1975. Approval for the study was granted by the NHS Research Ethics Committee London Camberwell St Giles. Consented participants were seen at the West London NHS Trust, where they underwent a range of clinical and cognitive assessments [25]. The cohort comprised cognitively healthy volunteers aged 40-59 years. The West London dataset was used here to avail of both baseline and follow-up testing data acquired for this cohort. 210 individuals (62 male; 148 female) were tested at baseline, with 188 (89.5%) (55 male; 133 female) retained at 2 years follow-up (Table 1).

**Table 1.**
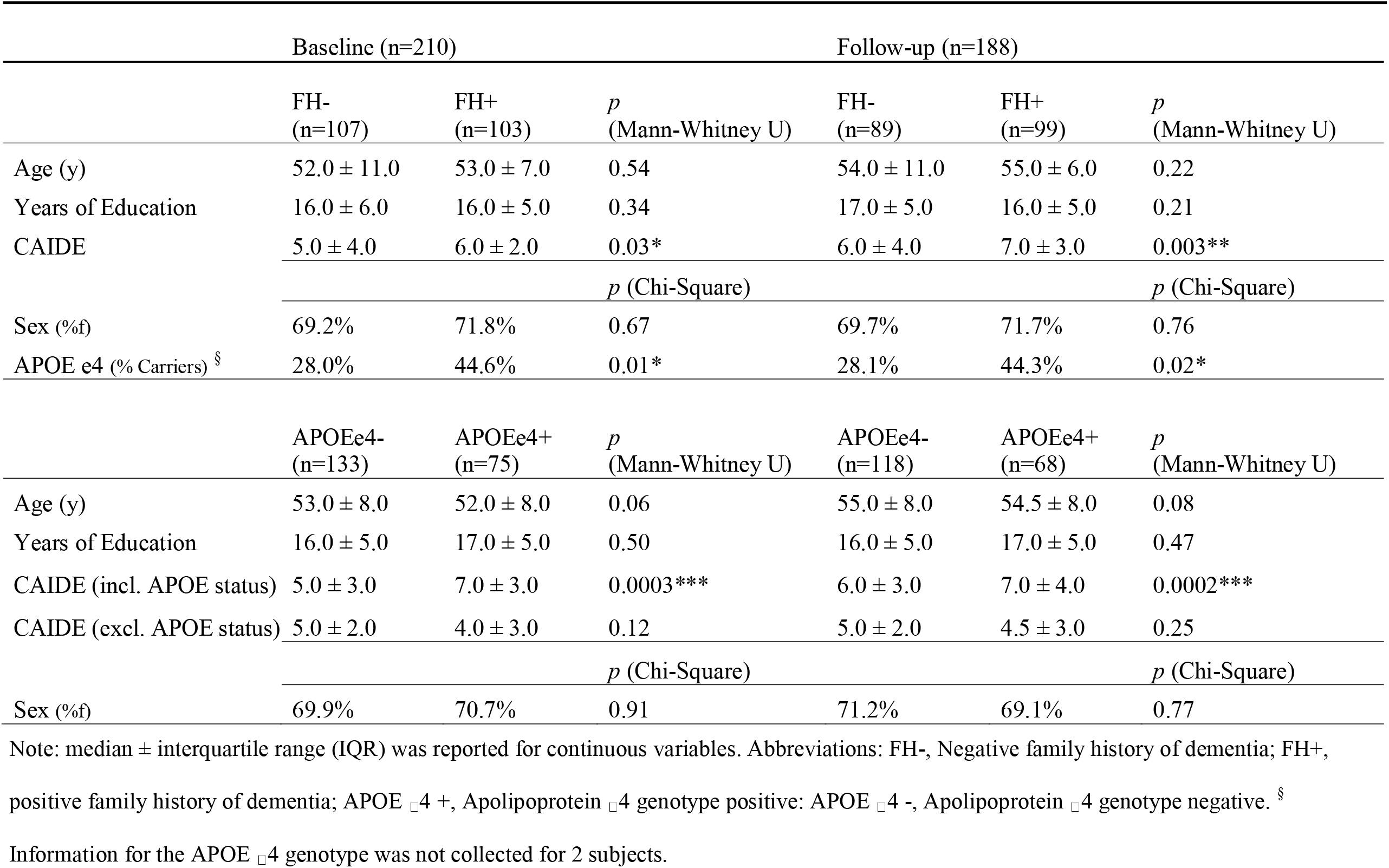
Demographic specifications of the cohort at baseline and follow-up based on the dementia family history and on APOE genotype

### Assessments

#### Risk factors

##### APOE_□_4 Genotyping

The process of APOE _□_4 allele identification is outlined in detail in Ritchie et al. (2017) [34]. In brief, genomic DNA was isolated from blood samples and APOE genotyping was performed. All members of the research and clinical teams were blind to the result of APOE genotyping. In this study, APOE _□_4 risk is determined by ≥1 APOE _□_4 allele. 75/210 carried ≥1 APOE _□_4 allele (See Table 1).

##### Family History

FH was determined by a ‘yes’/’no’ question during clinical visits, which asked participants whether a parent had a diagnosis of dementia. Participants were asked to include the dementia subtype if known, but answering ‘yes’ alone categorized a participant as FH+. The answer ‘no’ likely captured both participants with no family history of dementia, and participants for whom FH was unknown. In summary, participants were defined as FH+ if at least one parent was diagnosed with dementia. Cases where the FH was unknown or partially known were not recorded outside of the binary yes/no scoring. 103/210 were FH+.

##### Cardiovascular Risk Factors, Aging, and Incidence of Dementia (CAIDE) score

CAIDE is a composite scale of estimated future dementia risk based on mid-life cardiovascular measures [51-52]. It takes into consideration the individual’s age, sex, educational attainment, APOE _□_4 genotype, activity level, BMI, cholesterol and systolic blood pressure [23] and is scored on a range of 0 – 18. A higher score indicates greater risk. The CAIDE dementia risk score was calculated for each participant at baseline and follow-up.

#### Cognitive testing

The Addenbrooke’s Cognitive Assessment (ACE) III [53] was recorded at the follow-up session, but not at the baseline. Subsequent analyses that focused on the subset of assessments performed at both testing session did not include the ACE) III.

Cognitive function was assessed at baseline and follow up with the COGNITO neuropsychological battery [54], designed to examine information processing across a wide range of cognitive functions in adults of all ages and not restricted to those functions usually implicated in dementia detection in the elderly. Tests are administered using a tactile screen to capture information processing time as well as response accuracy and require about 40 minutes to complete. The tests, by order of presentation, are: reaction time; reading; comprehension of phonemes, phrases, and syntax; focused and divided attention in both visual and auditory modalities; visual working memory (visual tracking with auditory interference); the Stroop test; immediate, delayed, and recognition trials for verbal recall (name list); delayed recognition of spatial stimuli (faces); visuospatial associative learning; visuospatial span; form perception; denomination of common objects; spatial reasoning; copying of meaningful and meaningless figures; verbal fluency with se-mantic and phonetic prompts; immediate recall of a narrative; immediate recall of a description of the relative position of objects; vocabulary; implicit memory (recognition of new and previously learned material).

The COGNITO tests are designed to test several aspects of cognition, including attention (task: visual attention), memory (tasks: narrative recall, description recall, implicit memory, name-face association, working memory), language (tasks: phoneme comprehension, verbal fluency) and visuospatial abilities (task: geometric figure recognition) [54-55]. Based on previous studies [54-55], 11 summary variables from the COGNITO battery capturing the above functions were used here [for a list see Supporting Information (SI)].

Additionally, we used the Visual Short-Term Memory Binding task (VSTMBT) [56], a computer-based task that assesses visual short-term memory binding of single features, e.g., complex shape or color combinations, or feature conjunctions, e.g., shape and color combinations. In the single feature condition, participants must identify whether the test stimuli (three random 6-sided polygons) are the “same” as or “different” to the studied stimuli in terms of shape (shape only) or color (color only). In the binding condition, participants are required to correctly identify if both the shape and color of the test stimuli match studied stimuli. Two summary variables from the VSTMBT were the percentage of correctly recognized items from the two conditions.

#### Measurement of lifestyle activities

The Lifetime of Experiences Questionnaire (LEQ) [50], designed to take a lifespan approach to the measurement of cognitive resilience [57-59] and mental activity, measures engagement in a broad range of lifestyle activities across three stages of life: young adulthood (13-29 years), mid-life (30-64 years) and late life (65 years onwards). Therefore, the LEQ is preferable for looking at a mid-life cohort compared to other scales that capture dementia-specific risk related to modifiable lifestyle factors (e.g., LIBRA [60]). The LEQ comprises sub-scores capturing “specific” activities, reflecting the primary activity undertaken in each life stage and “non-specific” activities, reflecting engagement in physical, social and intellectual activities in any stage. For the purpose of this paper, we define mid-life ‘lifestyle’ as all the activities captured by the LEQ (below).

##### Mid-life specific score

The mid-life specific component score centers on occupation and comprises two sub-scores. For the first occupational sub-score, participants were asked to record their primary occupation in each 5-year interval from age 30 to age at assessment. Each reported occupation was scored according to the International Standard Classification of Occupations (ISCO 08) guidelines (https://www.ilo.org/public/english/bureau/stat/isco/isco08/), and scores were inverted and summed. The second sub-score is a measure of the managerial responsibility associated with reported occupations. If participants indicated that they were employed in a managerial capacity, the number of people that they oversaw in four of their reported occupations was documented. Managerial responsibility was scored as follows; 0 people = 8, 1-5 people = 16, 5-10 people = 24 and 10+ people = 32. The highest score is recorded as the managerial responsibility sub-score. The occupational history and managerial sub-scores were summed and multiplied by a normalization factor of 0.25. Normalization ensures that the mid-life specific and non-specific scores have comparable mean values [50].

##### Mid-life non-specific score

The non-specific score assesses frequency of engagement in 7 activities, capturing those of a physically, socially and intellectually stimulating nature, scored on a 5-point Likert scale of frequency (never, less than monthly, monthly, fortnightly, weekly, daily). Scores range from 0 – 35, with higher scores reflecting more frequent engagement in such activities. The items included in the scale are socializing with family or friends, practicing a musical instrument, practicing an artistic pastime, engagement in physical activity that is mildly, moderately, or vigorously energetic, reading, practicing a second language and travel. The travel item asks participants if they have visited any of a list of continents between the ages of 30-54. Responses were scored on a 5-point scale as follows; none, 1-2 regions, 3-4 regions, 5 regions, 6-7 regions, all regions.

### Statistical analyses

To reduce the number of multiple comparisons between the cognitive tests, given the multitude of cognitive assessments obtained for this cohort (13 summary variables, see SI and Table S1), in an independent study of this dataset [35], three composite cognitive components were extracted from the above cognitive assessments, by using rotated principal component analysis (rPCA) (Figure S1). In subsequent analyses, we used these cognitive components, rather than the individual tests to measure the impact of risk and protective lifestyle factors on cognition.

At baseline, the three components were: 1) C1: verbal, spatial and relational memory; 2) C2: working and short-term (single-feature) memory; 3) C3: verbal and visuospatial functions, and short-term (conjunctive) memory (Figure S1 A). Higher scores in C1 and C2 reflected better performance, whereas higher scores in C3 reflected poorer performance. For ease of visualization and comparability to C1 and C2, values of C3 were recoded with reversed sign.

The highest loading tasks and their weights for each component differed slightly for the cross-sectional and longitudinal data. There was high similarity between baseline and follow-up for C1 (*ϕ* = 0.90), and C3 (*ϕ* = 0.76), with relatively low similarly for C2 (*ϕ* = 0.40) (Figure S1 B). Although C1 and C3 had relatively high similarity across time, the longitudinal changes in these components could not be quantified by direct comparison, because they captured different cognitive functions at each time-point and any longitudinal change would be hard to interpret.

We used the Statistical Package for Social Sciences (SPSS V.26) and R software for all statistical analyses. The normality of the data was assessed by combining the visualization of a quantile-quantile plot and the Shapiro–Wilk test. Demographic and clinical information of the study cohort was analyzed across risk groups using chi-square (χ^*2*^ tests) for categorical variables and Mann Whitney U tests for continuous (discrete) variables, given that they were not normally distributed in this cohort. Subsequently, we used hierarchical regression models to look at the contribution of mid-life lifestyle factors (LEQ specific and non-specific scores) risk factors (APOE _□_4 genotype and FH) and of their interactions on cognitive performance, at baseline and follow-up. The effect of APOE _□_4 and CAIDE risk factors was modelled independently, in order to avoid modelling the variance associated with APOE genotype in the same model twice. In each case, dependent variables were the aforementioned composite cognitive components, each assessed in a separate model.

The variance of some parts of physical activity is accounted for twice in the model, in the CAIDE score and in the non-specific LEQ factor, resulting in weakening of its statistical contribution that can by captured by the lifestyle factor. Therefore, any effect of physical activity that can be captured by the lifestyle factor (independent variable) in the same model as CAIDE (independent variable), can only provide a conservative estimate of the contribution of physical activity to the dependent variable of cognitive performance.

As aforementioned, the baseline and follow-up datasets were analyzed independently, due to the slightly differing cognitive domains for each visit. Although the two datasets were not independent, we considered each of them in its own right, because during 2 years, a proportion of our participants may have had substantial brain health changes that are yet subthreshold to clinical manifestations. Therefore, the follow-up dataset has the potential to reveal the impact of age on variables of interest. Corrections for multiple comparisons were performed across all analyses.

To avoid multicollinearity, we mean centered continuous variables (specific and non-specific scores). Age, sex and years of education were included as covariates in hierarchical models. A significant interaction effect between a risk and a lifestyle factor would indicate that the effect of the lifestyle factor on cognitive performance differs across levels/values of the risk factor. For any observed interactions, we plotted the regression of the lifestyle factor on cognitive performance for each level/value of the risk factor [61], to interpret the effect. We then tested the significance of the slopes of the simple regression lines, to investigate in which level/value of the risk factor we found an effect of lifestyle on cognitive performance.

## Results

### Demographic characteristics

In the first PREVENT study phase, 210 participants were recruited from a single site. All 210 completed several cognitive tests and the LEQ questionnaire. 188 were also assessed at the follow up, two years later. At baseline, 2 participants were missing information relating to their APOE _□_4 genotype, with a further 2 missing cognitive data. The final baseline cohort that could be used for the analyses was N = 206. At follow-up, 12 participants were missing cognitive data, alongside the 2 participants who were missing APOE _□_4 genotype data. Therefore, the final follow-up cohort was N = 174. Mild cognitive impairment and dementia were ruled out based on detailed clinical assessment at baseline and follow-up. Please see the Supplementary Information and Table S1 for a list of all the outcome variables collected at baseline and follow-up.

Demographic specifications of the cohort at baseline and follow-up, stratified by APOE _□_4 genotype and family history of dementia, are shown in Table 1. There were no significant differences in age, sex or years of education between the groups. As expected, APOE _□_4 allele genotype was more frequently found in the FH+ than FH-group at baseline (p = 0.01) and follow up (p = 0.02). CAIDE scores (including APOE status) were significantly higher for the FH+ than FH-group at baseline (p = 0.03) and follow up (p = 0.003). Naturally, CAIDE scores including APOE status were significantly higher for the APOE _□_4+ than APOE _□_4-group at baseline (p = 0.0003) and follow up (p = 0.0002), but when APOE status was excluded the CAIDE scores did not differ between the two groups (Table 1).

### Verbal, Spatial and Relational memory

At baseline, a hierarchical regression model with lifestyle factors, i.e., the specific and non-specific LEQ scores, risk factors (APOE _□_4 and FH), and age, sex and years of education as covariates, and verbal, spatial and relational memory as the dependent variable was significant overall [F (7, 198) = 5.84, R^2^ = 0.17, p < 0.0001] (Supplementary Table 2a), and showed a significant positive association between verbal, spatial and relational memory and education [β (SE) = 0.09 (0.02), p < 0.0001] (Figure 1a). Similarly, at follow up the model’s performance was significant [F (7, 166) = 5.52, R^2^ = 0.19, p < 0.0001], and there was a significant positive association between verbal, spatial and relational memory and education [β (SE) = 0.10 (0.02), p < 0.0001] (Figure 1b). Higher education values were significantly associated with higher performance at both timepoints. The inclusion of risk by lifestyle interaction terms (FH × specific score, FH × non-specific score, APOE _□_4 × specific score, APOE _□_4 × non-specific score) into the hierarchical regression model (Supplementary Table 2b) did not show any significant associations between lifestyle x risk interaction terms and cognitive performance, at baseline or follow-up.

**Figure 1.**
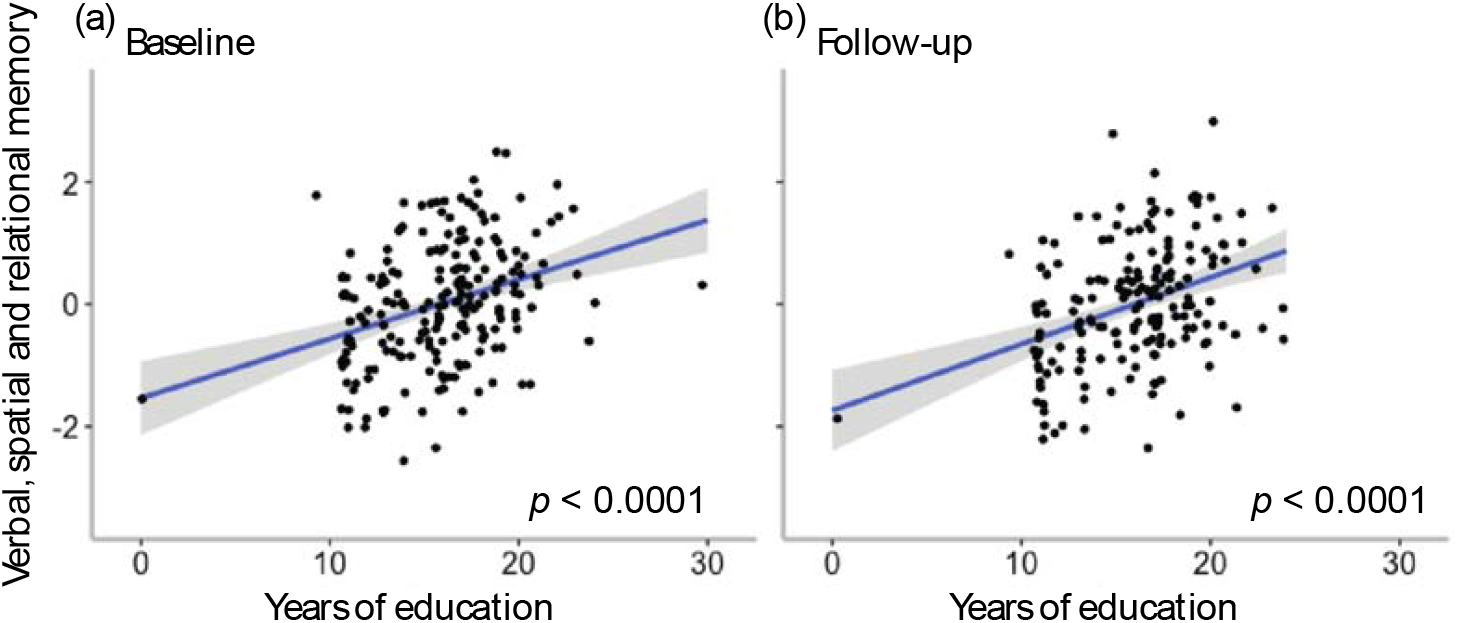
Association of years of education with verbal, spatial and relational memory performance at baseline (a) and follow-up (b). The x axis displays total reported years of education attained. On the y axis, higher scores represent better verbal, spatial and relational memory performance.

The hierarchical regression models with lifestyle factors and CAIDE as independent variables, and verbal, spatial and relational memory as the dependent was significant at baseline [F (3, 202) = 3.11, R^2^ = 0.04, p = 0.03] and at follow-up [F (3, 166) = 3.52, R^2^ = 0.04, p = 0.02] (Supplementary Table 2c). At both timepoints, we found a significant positive association between the non-specific LEQ factor and verbal, spatial and relational memory (baseline: [β (SE) = 0.04 (0.02), p = 0.02], figure 2a) and (follow up: [β (SE) = 0.07 (0.02), p = 0.002], figure 2b). More frequent engagement in physically, socially and intellectually stimulating activities was associated with better verbal, spatial and relational memory. This association was independent of CAIDE and therefore of age, sex and years of education, as included in this score. The inclusion of CAIDE by lifestyle interaction terms (CAIDE × specific score, CAIDE × non-specific score) into the hierarchical regression model (Supplementary Table 2d) did not show any significant associations between lifestyle x risk interaction terms and cognitive performance, at baseline or follow-up.

**Figure 2.**
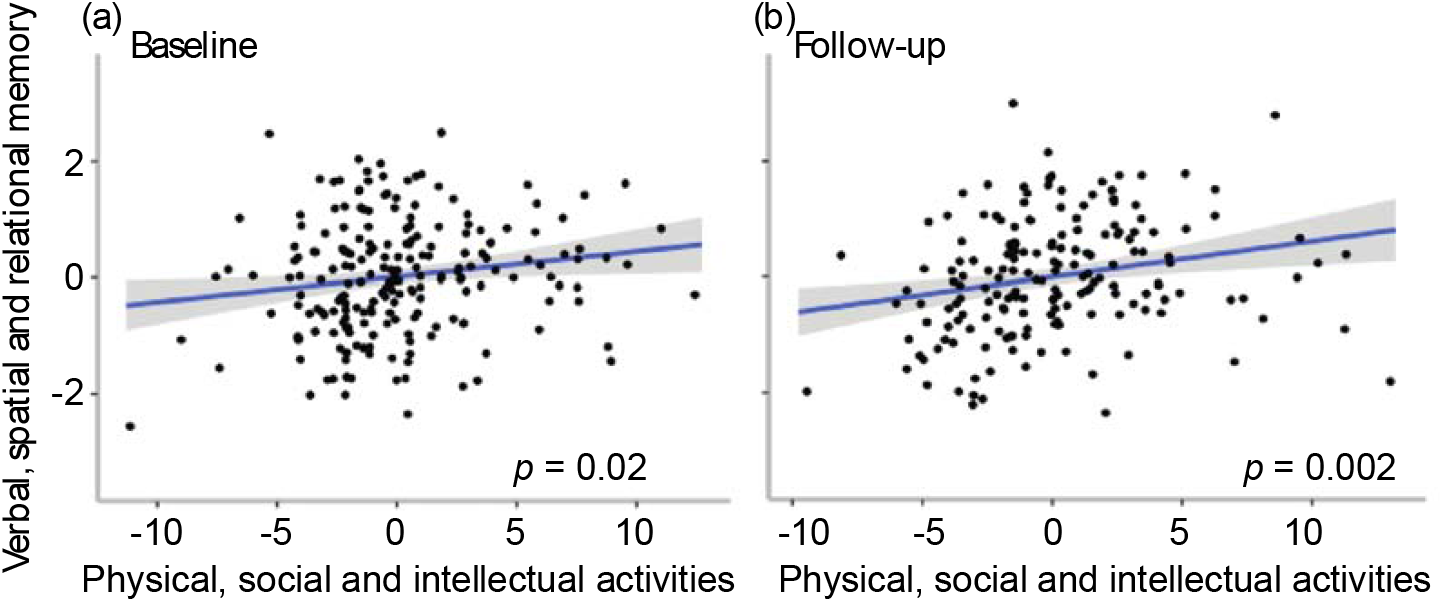
Association of physical, social and intellectual activities with verbal, spatial and relational memory performance at baseline (a) and follow-up (b). Physical, social and intellectual activities are mean-centred. On the x axis, higher scores represent more frequent engagement in physical, social and intellectual lifestyle activities, and on the y axis, higher scores represent better verbal, spatial and relational memory performance.

### Working and Short-Term (Single-Feature) Memory

Hierarchical regression analyses showed no significant associations of any of the independent variables and performance on the working and short-term (single-feature) memory, at baseline or follow up (Supplementary Table 3a, 3c) Furthermore, no significant associations with interaction terms were observed at either timepoint (Supplementary Table 3b, 3d).

Verbal and Visuospatial Functions, and Short-Term (Conjunctive) Memory At baseline, the hierarchical regression model was significant [F (7, 198) = 2.39, R^2^ = 0.08, p = 0.02], and showed a significant negative association between verbal and visuospatial function and short-term (conjunctive) memory with age [β (SE) = -0.04 (0.01), p = 0.006] (Supplementary Table 4a). Higher age was associated with poorer performance. At follow-up, there were no significant associations between the main independent variables with verbal and visuospatial function and short-term (conjunctive) memory. The inclusion of risk by lifestyle interaction terms in the hierarchical model revealed a significant association between the non-specific LEQ score x FH interaction term and cognitive performance [β (SE) = 0.11 (0.05), p = 0.01] (Supplementary Table 4b). To interpret this interaction, we investigated the relationship between cognitive performance and the non-specific LEQ score for the FH+ and FH-groups independently (Figure 3). We found a significant negative relationship between non-specific lifestyle activities and cognition in the FH+ group [β (SE)= 0.08 (0.03), p = 0.02), that was independent of age, sex, or years of education. We did not observe a relationship between non-specific lifestyle activities and cognition in the FH-group [β (SE) = -0.03 (0.03), p = 0.34]. These results suggested that for individuals with positive family history, more frequent engagement in the non-specific LEQ factor — namely physically, socially and intellectually engaging activities — was associated with better performance in verbal and visuospatial function and short-term (conjunctive) memory. Finally, we also observed trend associations between cognitive performance and the specific LEQ factor score x FH interaction term [β (SE) = -0.07 (0.04), p = 0.06]. This association, while suggestive of a role for occupational complexity on cognition, stratified by family history risk group, is weak and needs to be investigated further in future studies. Additionally, as previously reported [35] we observed a significant negative association between CAIDE and verbal and visuospatial functions and short-term (conjunctive) memory at baseline (Supplementary Table 4c). There were no associations between interactions of CAIDE x lifestyle and cognition at either baseline or follow up (Supplementary Table 4d).

**Figure 3.**
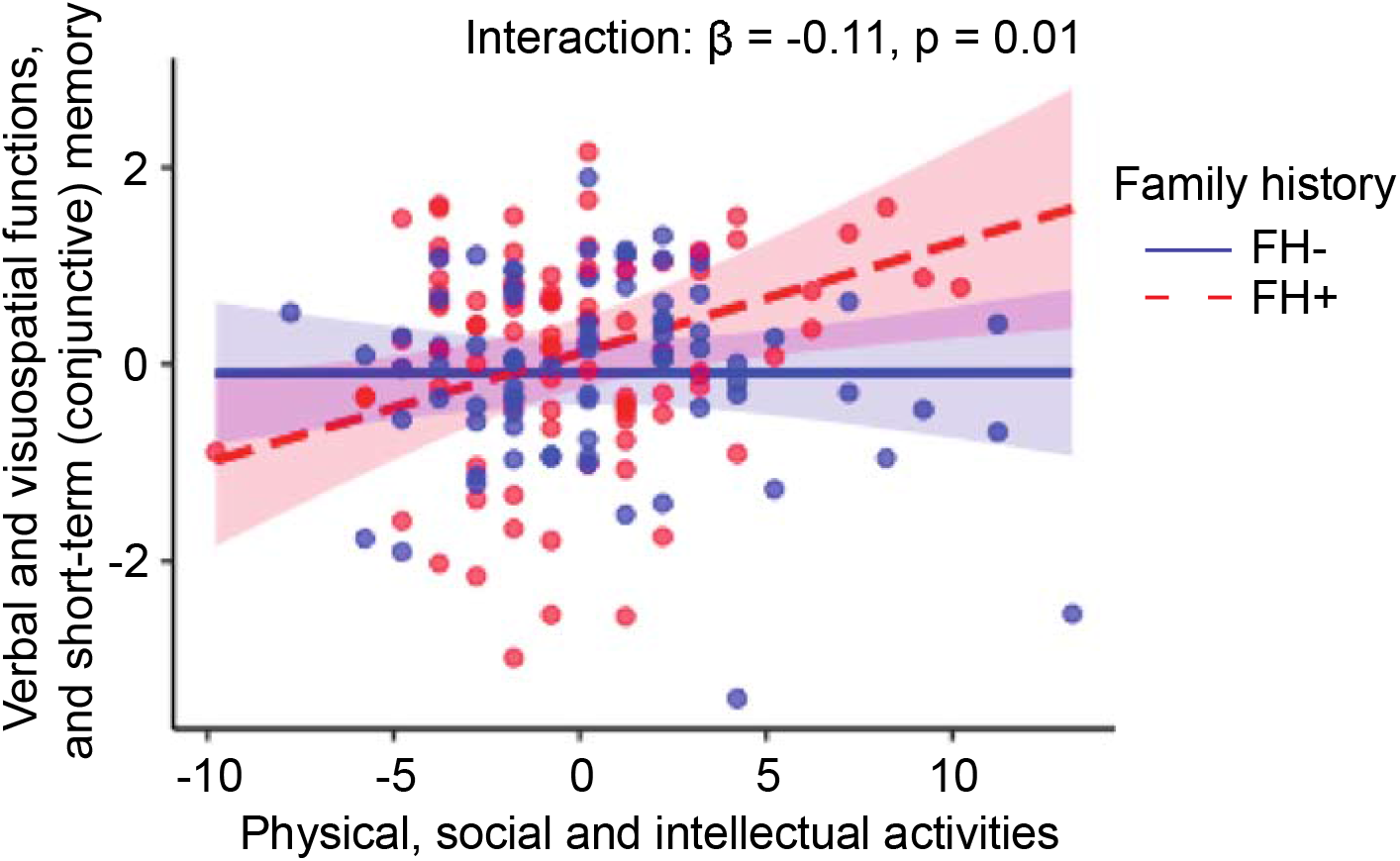
Interaction of physical, social and intellectual activities and family history of dementia on verbal and visuospatial functions, and short-term (conjunctive) memory at follow-up. On the x axis, higher scores represent more frequent engagement in physical, social and intellectual lifestyle activities, and on the y axis, higher scores represent poorer verbal and visuospatial functions, and short-term (conjunctive) memory. Physical, social and intellectual activities are mean-centred. The positive family history of dementia group showed a significant positive association between engagement in these activities and improved verbal and visuospatial functions, and short-term (conjunctive) memory. No significant association was seen for the negative family history group. Abbreviations: FH, family history; FH+, family history positive; FH-, family history negative.

## Conclusion

It is now acknowledged that Alzheimer’s Disease processes are present decades before the onset of clinical symptoms [2, 62, but, to date, it has remained unknown whether lifestyle factors can already protect against these early AD processes in mid-life. We asked whether these modifiable lifestyle activities impact cognition in middle-aged individuals who are cognitively healthy but at risk for late life AD. Lifestyle activities significantly impacted cognition in mid-life. Individuals with greater educational attainment showed stronger cognition in a composite dimension capturing verbal, spatial and relational memory. This result is consistent with a previous study [34] of this cohort showing that visuospatial abilities were positively associated with education. It is also consistent with epidemiological studies on older adults showing that education contributes to cognitive resilience in older life [48]. Our cohort is highly educated, with approximately 30% reaching a post-graduate qualification [34]. As the LEQ education measure captures total years of education, the effect of education we observed, reflects long-term effects set in motion from early life and young adulthood.

The key question in this study, however, was to investigate any additional contribution of activities undertaken in mid-life, independently of education. The first novel finding of this study was that more frequent engagement in physically, socially and intellectually stimulating activities in mid-life was associated with stronger cognition in a composite dimension capturing verbal, spatial and relational memory, both at baseline and at follow-up. This effect was independent of sex, age, years of education and cardiovascular factors captured by the CAIDE score. The second novel finding was that physically, socially and intellectually stimulating activities undertaken in mid-life had a significant effect in the cognition of middle-aged individuals at risk for late-life AD. Specifically, higher engagement in these activities was associated with significantly stronger cognition in another composite dimension, capturing verbal and visuospatial functions and short-term (conjunctive) memory in cognitively healthy individuals, who were at risk for late life AD though family history of dementia, at follow up. Importantly, this effect was independent of age, sex, and years of education. The presence of this effect in the follow-up but not the baseline dataset is likely due to the older age of the cohort at follow-up. Short-term (conjunctive) memory functions have been found impaired in the pre-symptomatic stages of AD [56]. Additionally, impaired visuospatial function is one of the earliest cognitive deficits observed in AD [63-64] and has previously been linked to increased AD risk in this cohort [34-35]. In similarly cognitively healthy, but older (> 65 years) North American cohorts [65-66], changes in cerebrospinal fluid biomarkers related to inflammation have been associated with changes in visuospatial cognitive performance, thereby suggesting a biochemically-mediated effect of early pathology in presymptomatic individuals.

Taken together, our results suggest that stimulating lifestyle activities may boost cognitive functions that are very vulnerable to AD risk and early AD neuropathology [56, 63-64]. However, we caution that the interpretability of the effect of lifestyle activities on each individual function is limited by their composite assessment in this study and requires individuation in future studies with a longer longitudinal follow-up window. Physically, socially and intellectually stimulating mid-life activities have been shown to influence late-life cognition, suggesting that they contribute to the cognitive resilience that mitigates the effect of age-related cognitive decline and AD neuropathology [40, 49]. Our finding advances understanding by showing that engagement with these activities contributes to cognitive resilience to risk of AD, or even incipient AD neuropathology, from mid-life, in individuals who are presently cognitively healthy.

Family history of dementia is a well-established risk factor [16, 19-20], independent of the genetic risk bestowed by APOE _□_4 genotype, that captures both genetic and environmental risk influences. For example, an individual who has one or more parent with dementia, may, independently to the presence of the APOE _□_4 genotype, be exposed to negative environmental influences that contribute to neuropathology, including increased stress [67], caregiver burden [68-69] and reduced participation in enriching environments due to caregiving duties [70]. Therefore, alongside contributing to cognitive resilience, enhanced engagement with physically, socially and intellectually stimulating activities may positively impact individuals with a family history of dementia by counteracting these negative environmental influences.

What might be the mechanism by which stimulating activities impact cognition in midlife? Models of AD disease delineated by neuropathological staging [9] place the locus coeruleus, a small nucleus in the pontine tegmentum region of the brainstem, at the pathogenesis of AD [10-11, 71]. The LC is responsible for the production of neurotransmitter noradrenaline (NA) [72], a major driver of the brain’s arousal system, which strongly modulates high-order cognition. Novelty provides a key trigger for arousal, and, thus, the LC–NA activity occurs strongly in response to novel stimuli. Therefore, stimulating activities upregulate the LC and may protect it from early neurodegeneration due to pTau deposition. Furthermore, as the brain site of noradrenaline, the LC is key to the protective effect of lifestyle factors on the whole brain. Studies suggests that environmental enrichment, through activities such as those measured in this study, upregulates the noradrenergic system, which otherwise depletes with age [47] and AD pathology [10], leading to compensatory brain mechanisms for cognitive function, such as the strengthening of the fronto-parietal brain and other large-scale brain networks [45-46].

In summary, our findings suggest that modifiable lifestyle activities may offset AD risk-related cognitive decrements in mid-life and support the targeting of certain modifiable lifestyle activities for the prevention of Alzheimer’s Disease, especially in those with a family history. These activities include socializing with family or friends, practicing a musical instrument, practicing an artistic pastime, engagement in energetic physical activities, reading, practicing a second language and travel. Given the apparent lifestyle contributions both as risk and protective factors of dementia [4], a healthy lifestyle may be the individual’s current best defense against sporadic late onset AD. The modifiability of the lifestyle activities identified here renders them promising cost-effective candidates for intervention and prevention strategies from early life. These activities may be particularly significant for non-pharmacological interventions for AD in low and middle-income countries (LMIC), where barriers to education are more prevalent than in high income countries [4].

### Methodological considerations

As education is strongly positively linked to IQ [73] and our cohort was highly educated, the question of reverse causation arises [40, 49, 74]. This points to the possibility that cognitive abilities may determine engagement in stimulating activities, rather than the inverse. However, the effect on mid-life verbal and visuospatial functions, and short-term (conjunctive) memory is independent of the total years of education, which shows that education does not directly drive this effect. Furthermore, we found that mid-life lifestyle activities were associated with improved cognitive performance only in the family history positive group, which is not, at least not prima facie, more educated than the family history negative group. Thus, this effect is likely independent of any indirect effects of education.

The use of composite cognitive domains that capture slightly different functions in the two assessment points [35] prevents investigation of the impact of lifestyle on cognitive changes over time in this study. Nevertheless, previous studies from this cohort have shown only subtle changes over the two-year period [75-76], possibly due to the relatively young age range of the sample and the short follow up window [34, 55]. Therefore, future studies that follow this cohort over a longer period and test hypotheses informed by the previous study waves are needed to determine the longitudinal impact of lifestyle activities in cognitively healthy middle-aged individuals at risk for late life AD.

It is worth noting that we found no evidence that lifestyle factors can mitigate the impact of the CAIDE risk, or for interactions between CAIDE, lifestyle and cognition, as reported in some previous studies (see Kivipelto et al. 2018 [77] for a review). Unlike the CAIDE score (including systolic blood pressure, BMI, total cholesterol, physical activity), the lifestyle factors captured by our instrument (the LEQ) are not primarily related to cardiovascular wellbeing (with the exception of exercise, 1/9 items), and therefore not likely to impact greatly on cardiovascular health in this relatively young cohort (i.e., 40–59 years). By contrast, the studies reviewed by Kivipelto et al. (2018) include older populations (> 60–70 years), where cardiovascular factors are more prevalent drivers of health vulnerabilities, and furthermore those studies, unlike the present one, had a strong focus on cardiovascular health, including nutritional guidance, exercise and monitoring and management of metabolic and vascular risk factors.

## Supporting information

SupplementaryInformation_Heneghan_Deng_etal

## Data Availability

All data produced in the present study are available upon reasonable request to the authors.

## Acknowledgement

F.D. was funded by the Provost PhD Award Scheme from Trinity College Dublin, to L.N. L.N. was also funded by a L’Oréal-UNESCO for Women In Science International Rising Talent Award, the Welcome Trust Institutional Strategic Support grant, and the Global Brain Health Institute Project Grant.

The PREVENT-Dementia study is supported by the UK Alzheimer’s Society (grant numbers 178 and 264), the US Alzheimer’s Association (grant number TriBEKa-17-519007) and philanthropic donations.

We thank all PREVENT-Dementia participants for their enthusiastic participation in this study. We also thank the Research Delivery service at West London NHS Trust and the Wolfson Clinical Imaging Facility at Imperial College London for their support in running the study.

## Conflict of interest

The authors declare no conflict of interest.

## Notes

### Competing Interest Statement

The authors have declared no competing interest.

### Funding Statement

This study was funded by the UK Alzheimers Society (grant numbers 178 and 264), the US Alzheimers Association (grant number TriBEKa-17-519007) and philanthropic donations.

### Author Declarations

NHS Research Ethics Committee of London Camberwell St Giles gave ethical approval for this work.

### Summary of Updates

Manuscript updated to revise and clarify on comments raised by reviewers. Tables in supplementary materials updated to include more detail on cognitive variables assessed.

