## SupplementaryInformation_Heneghan_Deng_etal for "Modifiable lifestyle activities affect cognition in cognitively healthy middle-aged individuals at risk for late-life Alzheimer’s Disease"

**Title**

**Running title:** Midlife lifestyle, AD risk and cognition

**Authors**

Amy **Heneghan** ^ab1^, Feng **Deng** ^ab1^, Katie **Wells** ^c^, Karen **Ritchie** ^d^, Graciela **Muniz-Terrera** ^e^, Craig W **Ritchie** ^f^, Brian **Lawlor** ^ab^, Lorina **Naci** ^ab*^

^a^ Trinity College Institute of Neuroscience, School of Psychology, Trinity College Dublin, Dublin, Ireland.

^b^ Global Brain Health Institute, Trinity College Dublin, Dublin, Ireland.

^c^ Edinburgh Dementia Prevention, University of Edinburgh, Edinburgh, UK.

^d^ INSERM and University of Montpellier, Montpellier, France.

^1^ These authors contributed equally to this work

^*^ **Corresponding author:**

Lorina Naci

School of Psychology

Trinity College Institute of Neuroscience

Global Brain Health Institute

Trinity College Dublin

Dublin, Ireland

**Materials and Methods**

The eleven cognitive summary variables from the COGNITO battery were:

1. Working memory: the simultaneous presentation of auditory and visual attention tasks assessed by subtracting the time taken in milliseconds on this double task from the visual task alone.
2. Working memory: the simultaneous presentation of auditory and visual attention tasks assessed by total number of correct answers for visual form recognition on this double task.
3. Narrative recall: total number of correct elements on immediate recall of a story with a temporal progression requiring attention to macrostructure.
4. Description recall: total number of correct elements recalled of a description without thematic progression requiring attention to microstructure and recall of spatial location. The narrative and description recall are similar in terms of word frequency in the language and syntactic structure.
5. Implicit memory: difference in the number of steps in the progressive build-up of names on the screen required for recognition between names never seen and number of names previously learnt in an immediate recall task.
6. Name-face association: number of faces recognized after a delay from a series of 18 faces of which 9 have been previously shown with their corresponding names.
7. Form perception: number of correct answers in the matching of complex forms to a multiple-choice array.
8. Form perception speed: mean time taken in milliseconds for each trial.
9. Phoneme comprehension: number of correct responses in the matching of a word with an image presented as part of a multiple-choice array including semantic, morphological and phonetic distractors.
10. Phoneme comprehension speed: mean time taken in milliseconds to perform.
11. Verbal fluency: total sum of the number of words generated in 30s using both a semantic (vegetables) and phonemic (letter P) cue.

The two summary variables from the Visual Short-term Memory Binding task were:

12. Visual short-term memory binding test (shape only condition): percentage of correct recognition of the shape of presented visual stimuli after a short period of retention.
13. Visual short-term memory binding test (shape-color binding condition): percentage of correct recognition of combinations of shape and color of presented visual stimuli after a short period of retention.

**Figures and Legends**

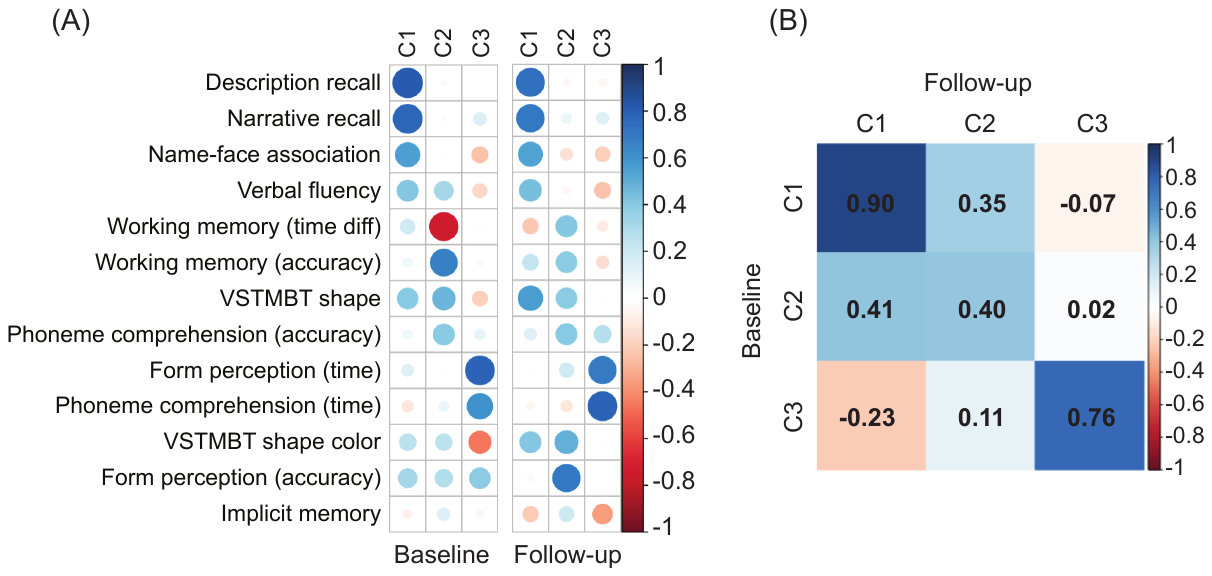

Supplementary Figure 1. Component loadings and similarity between baseline and follow-up. (A) The loading of each cognitive measure (in rows) on each cognitive component (in columns). The color bar indicates loading values, higher loading values indicate stronger contribution to corresponding components. (B) Similarity of components between baseline and follow-up. Higher values in the color bar indicate higher similarity across testing sessions. Abbreviations: C, component; C1, verbal, spatial and relational memory; C2, Working and short-term (single feature) memory; C3, verbal and visuospatial functions, and short-term (conjunctive) memory; VSTMBT, visual short-term memory binding test; diff, difference. Adapted from Deng et al., 2022.

**Tables**

Supplementary Table 1. Complete list of risk factors and tests obtained at baseline and follow-up

| Baseline (N=210) | Follow-up (N=188) |
| --- | --- |
| **Risk factors** | **Risk factors** |
| *Family history* | *Family history* |
| *APOE _Ɛ_4* (Missing = 2) | *APOE _Ɛ_4 (*Missing = 2) |
| *CAIDE (*Missing = 2) | *CAIDE (*Missing = 6) |
| Age | Age |
| Sex | Sex |
| Years of education | Years of education |
| Systolic blood pressure | Systolic blood pressure |
| BMI | BMI (Missing = 1) |
| Cholesterol | Cholesterol (Missing = 2) |
| Physical activity | Physical activity (Missing = 1) |
| APOE _Ɛ_4 (Missing = 2) | APOE _Ɛ_4 (Missing = 2) |
| **Neuropsychological assessments** | **Neuropsychological assessments** |
| *COGNITO* | *COGNITO* (Missing = 4) |
| Working memory (the simultaneous presentation of auditory and visual attention tasks assessed by subtracting the time taken in milliseconds on this double task from the visual task alone) | Working memory (the simultaneous presentation of auditory and visual attention tasks assessed by subtracting the time taken in milliseconds on this double task from the visual task alone) |
| Working memory (the simultaneous presentation of auditory and visual attention tasks assessed by total number of correct answers for visual form recognition on this double task) | Working memory (the simultaneous presentation of auditory and visual attention tasks assessed by total number of correct answers for visual form recognition on this double task) |
| Narrative recall (total number of correct elements on immediate recall of a story with a temporal progression requiring attention to macrostructure) | Narrative recall (total number of correct elements on immediate recall of a story with a temporal progression requiring attention to macrostructure) |
| Description recall (total number of correct elements recalled of a description without thematic progression requiring attention to microstructure and recall of spatial location. The narrative and description recall are similar in terms of word frequency in the language and syntactic structure) | Description recall (total number of correct elements recalled of a description without thematic progression requiring attention to microstructure and recall of spatial location. The narrative and description recall are similar in terms of word frequency in the language and syntactic structure) |
| Implicit memory (difference in the number of steps in the progressive build-up of names on the screen required for recognition between names never seen and number of names previously learnt in an immediate recall task) | Implicit memory (difference in the number of steps in the progressive build-up of names on the screen required for recognition between names never seen and number of names previously learnt in an immediate recall task) |
| Name-face association (number of faces recognized after a delay from a series of 18 faces of which 9 have been previously shown with their corresponding names) | Name-face association (number of faces recognized after a delay from a series of 18 faces of which 9 have been previously shown with their corresponding names) |
| Form perception: number of correct answers in the matching of complex forms to a multiple-choice array | Form perception: number of correct answers in the matching of complex forms to a multiple-choice array |
| Form perception speed: mean time taken in milliseconds for each trial | Form perception speed: mean time taken in milliseconds for each trial |
| Phoneme comprehension (number of correct responses in the matching of a word with an image presented as part of a multiple-choice array including semantic, morphological and phonetic distractors) | Phoneme comprehension (number of correct responses in the matching of a word with an image presented as part of a multiple-choice array including semantic, morphological and phonetic distractors) |
| Phoneme comprehension speed (mean time taken in milliseconds to perform) | Phoneme comprehension speed (mean time taken in milliseconds to perform) |
| Verbal fluency (total sum of the number of words generated in 30s using both a semantic (vegetables) and phonemic (letter P) cue) | Verbal fluency (total sum of the number of words generated in 30s using both a semantic (vegetables) and phonemic (letter P) cue) |
| *VSTMBT (*Missing = 2) | *VSTMBT (*Missing = 11) |
|  | *ACEIII* (Missing = 3) |
| **Lifestyle questionnaire** | **Lifestyle questionnaire** |

Supplementary Table 2a. FH & APOE- Regression coefficients for Verbal, Spatial and Relational Memory (Component 1) at Baseline & Follow-up

|  |  | Baseline | | | Follow-up | | |
| --- | --- | --- | --- | --- | --- | --- | --- |
| Model summary | | R^2^ | F | p | R^2^ | F | p |
|  |  | 0.17 | 5.84 | <0.0001 | 0.19 | 5.52 | <0.0001 |
| DV | IV | β (SE) | | p | β (SE) | | p |
| Component 1 | Specific | -0.02(0.02) | | 0.36 | -0.02 (0.02) | | 0.30 |
|  | Non-specific | 0.03 (0.02) | | 0.18 | 0.03 (0.02) | | 0.11 |
|  | Family history | 0.07 (0.13) | | 0.61 | -0.12 (0.15) | | 0.41 |
|  | APOE_Ɛ_4 | 0.26 (0.14) | | 0.06 | 0.27 (0.15) | | 0.07 |
|  | Age | -0.02 (0.01) | | 0.27 | -0.005 (0.02) | | 0.74 |
|  | Sex | 0.24 (0.14) | | 0.09 | 0.23 (0.16) | | 0.15 |
|  | Years of education | 0.09 (0.02) | | <0.0001 | 0.10 (0.02) | | <0.0001 |

DV, dependent variable; IV, independent variable; APOE _Ɛ_4, Apolipoprotein _Ɛ_4.

Supplementary Table 2b. FH & APOE- Regression coefficients for Verbal, Spatial and Relational Memory (Component 1) at Baseline & Follow-up including interaction terms

|  |  | Baseline | | | Follow-up | | | |
| --- | --- | --- | --- | --- | --- | --- | --- | --- |
|  | Model summary | R^2^ | F | p | R^2^ | | F | p |
|  |  | 0.18 | 3.88 | <0.0001 | 0.20 | | 3.60 | 0.0001 |
| DV | IV | β (SE) | | p | β (SE) | | | p |
| Component 1 | Specific | -0.01 (0.02) | | 0.57 | -0.04 (0.03) | | | 0.17 |
|  | Non-specific | 0.01 (0.03) | | 0.67 | 0.02 (0.03) | | | 0.49 |
|  | Family history | 0.06 (0.13) | | 0.63 | -0.13 (0.15) | | | 0.39 |
|  | APOE_Ɛ_4 | 0.25 (0.14) | | 0.07 | 0.26 (0.15) | | | 0.09 |
|  | FH*Specific | -0.01 (0.03) | | 0.86 | 0.03 (0.03) | | | 0.44 |
|  | APOE_Ɛ_4*Specific | 0.01 (0.03) | | 0.70 | 0.02 (0.04) | | | 0.68 |
|  | FH*Non-specific | -0.02 (0.04) | | 0.60 | 0.004 (0.04) | | | 0.92 |
|  | APOE_Ɛ_4* Non-specific | 0.05 (0.04) | | 0.18 | 0.02 (0.04) | | | 0.57 |
|  | Age | -0.02 (0.01) | | 0.19 | -0.005 (0.02) | | | 0.75 |
|  | Sex | 0.21 (0.15) | | 0.16 | 0.22 (0.16) | | | 0.17 |
|  | Years of education | 0.09 (0.02) | | <0.0001 | 0.09 (0.02) | | | <0.0001 |

DV, dependent variable; IV, independent variable; APOE _Ɛ_4, Apolipoprotein _Ɛ_4; FH, family history.

Supplementary Table 2c. CAIDE - Regression coefficient for Verbal, Spatial and Relational Memory (Component 1) at Baseline & Follow-up

|  |  | Baseline | | | | | Follow-up | | | | |
| --- | --- | --- | --- | --- | --- | --- | --- | --- | --- | --- | --- |
|  | Model summary | R^2^ | | F | p | | R^2^ | | F | | p |
|  |  | 0.04 | | 3.11 | 0.03 | | 0.06 | | 3.52 | | 0.02 |
| DV | IV | β (SE) | | | p | | β (SE) | | | | p |
| Component 1 | CAIDE | -0.05 (0.03) | | | 0.10 | | -0.01 (0.03) | | | | 0.78 |
|  | Specific | -0.01 (0.02) | | | 0.64 | | -0.02 (0.02) | | | | 0.36 |
|  | Non-Specific | 0.04 (0.02) | | | 0.02 | | 0.07 (0.02) | | | | 0.002 |

DV, dependent variable; IV, independent variable; CAIDE, Cardiovascular Aging and Dementia risk score.

Supplementary Table 2d. CAIDE - Regression coefficients for Verbal, Spatial and Relational Memory (Component 1) at Baseline & Follow-up including interaction terms.

|  |  | Baseline | | | | | | Follow-up | | | | |
| --- | --- | --- | --- | --- | --- | --- | --- | --- | --- | --- | --- | --- |
| Model summary | | | | R^2^ | F | p | | R^2^ | | F | | p |
|  |  |  |  | 0.05 | 2.04 | 0.07 | | 0.08 | | 2.76 | | 0.02 |
| DV | IV | β (SE) | | | | p | | β (SE) | | | | p |
| Component 1 | CAIDE | -0.05 (0.03) | | | | 0.11 | | 0.01 (0.03) | | | | 0.78 |
|  | Specific | -0.01 (0.02) | | | | 0.71 | | -0.02 (0.02) | | | | 0.32 |
|  | Non-Specific | 0.04 (0.02) | | | | 0.02 | | 0.06 (0.02) | | | | 0.003 |
|  | CAIDE*Specific | 0.01 (0.01) | | | | 0.34 | | 0.01 (0.01) | | | | 0.11 |
|  | CAIDE*Non-specific | -0.001 (0.01) | | | | 0.87 | | 0.005 (0.01) | | | | 0.60 |

DV, dependent variable; IV, independent variable; CAIDE, Cardiovascular Aging and Dementia risk score.

Supplementary Table 3a. FH & APOE - Regression coefficients for Working and Short-Term (single-feature) Memory (Component 2) at Baseline & Follow-up

|  |  | Baseline | | | Follow-up | | |
| --- | --- | --- | --- | --- | --- | --- | --- |
|  | Model summary | R^2^ | F | p | R^2^ | F | p |
|  |  | 0.05 | 1.44 | 0.19 | 0.05 | 1.36 | 0.23 |
| DV | IV | β (SE) | | p | β (SE) | | p |
| Component 2 | Specific | 0.02 (0.02) | | 0.22 | 0.01 (0.02) | | 0.57 |
|  | Non-specific | 0.01 (0.02) | | 0.78 | -0.03 (0.02) | | 0.17 |
|  | Family history | 0.08 (0.14) | | 0.59 | -0.02 (0.16) | | 0.89 |
|  | APOE_Ɛ_4 | <-0.001 (0.15) | | 1.00 | 0.22 (0.16) | | 0.19 |
|  | Age | -0.02 (0.02) | | 0.15 | -0.02 (0.02) | | 0.23 |
|  | Sex | -0.20 (0.16) | | 0.21 | -0.29 (0.17) | | 0.09 |
|  | Years of education | 0.04 (0.02) | | 0.10 | -0.003 (0.02) | | 0.90 |

DV, dependent variable: IV, independent variable; APOE _Ɛ_4, Apolipoprotein _Ɛ_4.

Supplementary Table 3b. FH & APOE - Regression coefficients for Working and Short-Term (single-feature) Memory (Component 2) at Baseline & Follow-up including interaction terms

|  | |  | Baseline | | | Follow-up | | |
| --- | --- | --- | --- | --- | --- | --- | --- | --- |
|  | | Model summary | R^2^ | F | p | R^2^ | F | p |
|  | |  | 0.06 | 1.08 | 0.38 | 0.08 | 1.29 | 0.23 |
| DV | IV | | β (SE) | | p | β (SE) | | p |
| Component 2 | Specific | | 0.03 (0.03) | | 0.27 | 0.01 (0.03) | | 0.81 |
|  | Non-specific | | 0.02 (0.03) | | 0.51 | -0.07 (0.03) | | 0.04 |
|  | Family history | | 0.08 (0.14) | | 0.59 | -0.03 (0.16) | | 0.87 |
|  | APOE_Ɛ_4 | | 0.01 (0.15) | | 0.93 | 0.19 (0.16) | | 0.24 |
|  | FH*Specific | | 0.003 (0.03) | | 0.94 | 0.01 (0.04) | | 0.80 |
|  | APOE_Ɛ_4*Specific | | -0.03 (0.04) | | 0.35 | -0.01 (0.04) | | 0.83 |
|  | FH*Non-specific | | 0.002 (0.04) | | 0.97 | 0.09 (0.05) | | 0.06 |
|  | APOE_Ɛ_4* Non-specific | | -0.04 (0.04) | | 0.39 | -0.001 (0.05) | | 0.99 |
|  | Age | | -0.02 (0.02) | | 0.21 | -0.02 (0.02) | | 0.30 |
|  | Sex | | -0.16 (0.16) | | 0.31 | -0.33 (0.17) | | 0.06 |
|  | Years of education | | 0.04 (0.02) | | 0.12 | -0.004 (0.02) | | 0.86 |

Note: DV, dependent variable; IV, independent variable; APOE _Ɛ_4, Apolipoprotein _Ɛ_4.

Supplementary Table 3c. CAIDE - Regression coefficient for Working and Short-Term (single-feature) Memory (Component 2) at Baseline & Follow-up

|  |  | Baseline | | | Follow-up | | |
| --- | --- | --- | --- | --- | --- | --- | --- |
|  | Model summary | R^2^ | F | p | R^2^ | F | p |
|  |  | 0.02 | 1.25 | 0.29 | 0.02 | 0.89 | 0.45 |
| DV | IV | β (SE) | | p | β (SE) | | p |
| Component 2 | CAIDE | -0.03 (0.03) | | 0.25 | -0.001 (0.03) | | 0.98 |
|  | Specific | 0.03 (0.02) | | 0.11 | 0.01 (0.02) | | 0.63 |
|  | Non-Specific | 0.01 (0.02) | | 0.57 | -0.04 (0.02) | | 0.11 |

DV, dependent variable; IV, independent variable; CAIDE, Cardiovascular Aging and Dementia risk score.

Supplementary Table 3d. CAIDE - Regression coefficient for Working and Short-Term (single-feature) Memory (Component 2) at Baseline & Follow-up including interaction terms.

|  |  | Baseline | | | Follow-up | | | |
| --- | --- | --- | --- | --- | --- | --- | --- | --- |
|  | Model summary | R^2^ | F | p | | R^2^ | F | p |
|  |  | 0.03 | 1.07 | 0.38 | | 0.02 | 0.60 | 0.70 |
| DV | IV | β (SE) | | p | | β (SE) | | p |
| Component 2 | CAIDE | -0.03 (0.03) | | 0.34 | | 0.001 (0.03) | | 0.99 |
|  | Specific | 0.03 (0.02) | | 0.10 | | 0.01 (0.02) | | 0.62 |
|  | Non-Specific | 0.01 (0.02) | | 0.63 | | -0.04 (0.02) | | 0.12 |
|  | CAIDE*Specific | 0.01 (0.01) | | 0.33 | | 0.004 (0.01) | | 0.61 |
|  | CAIDE*Non-specific | 0.01 (0.01) | | 0.51 | | -0.003 (0.01) | | 0.71 |

DV, dependent variable; IV, independent variable; CAIDE, Cardiovascular Aging and Dementia risk score.

Supplementary Table 4a. FH & APOE- Regression coefficient values for Verbal and Visuospatial Functions, and Short-Term (conjunctive) Memory (Component 3) at Baseline & Follow-up

|  |  | Baseline | | | Follow-up | | |
| --- | --- | --- | --- | --- | --- | --- | --- |
|  | Model summary | R^2^ | F | p | R^2^ | F | p |
|  |  | 0.08 | 2.39 | 0.02 | 0.04 | 0.91 | 0.50 |
| DV | IV | β (SE) | | p | β (SE) | | p |
| Component 3 | Specific | 0.001 (0.02) | | 0.94 | 0.004 (0.02) | | 0.83 |
|  | Non-specific | 0.03 (0.02) | | 0.10 | 0.02 (0.02) | | 0.51 |
|  | Family history | -0.21 (0.14) | | 0.13 | 0.20 (0.16) | | 0.20 |
|  | APOE_Ɛ_4 | -0.09 (0.15) | | 0.53 | -0.19 (0.16) | | 0.25 |
|  | Age | -0.04 (0.01) | | 0.006 | -0.03 (0.02) | | 0.11 |
|  | Sex | 0.22 (0.15) | | 0.16 | 0.18 (0.17) | | 0.29 |
|  | Years of education | -0.01 (0.02) | | 0.55 | 0.01 (0.02) | | 0.77 |

Note: DV, dependent variable; IV, independent variable; APOE _Ɛ_4, Apolipoprotein _Ɛ_4.

Supplementary Table 4b. FH & APOE- Regression coefficient values for Verbal and Visuospatial Functions, and Short-Term (conjunctive) Memory (Component 3) at Baseline & Follow-up including interaction terms

|  |  | Baseline | | | Follow-up | | |
| --- | --- | --- | --- | --- | --- | --- | --- |
|  | Model summary | R^2^ | F | p | R^2^ | F | p |
|  |  | 0.09 | 1.76 | 0.06 | 0.09 | 1.48 | 0.14 |
| DV | IV | β (SE) | | p | β (SE) | | p |
| Component 3 | Specific | 0.01 (0.03) | | 0.75 | 0.01 (0.03) | | 0.63 |
|  | Non-specific | 0.05 (0.03) | | 0.07 | 0.0004 (0.03) | | 0.99 |
|  | Family history | -0.22 (0.14) | | 0.12 | 0.20 (0.16) | | 0.20 |
|  | APOE_Ɛ_4 | -0.08 (0.15) | | 0.61 | -0.19 (0.16) | | 0.24 |
|  | FH*Specific | 0.004 (0.03) | | 0.90 | -0.07 (0.04) | | 0.06 |
|  | APOE_Ɛ_4*Specific | -0.03 (0.04) | | 0.36 | 0.06 (0.04) | | 0.09 |
|  | FH*Non-specific | -0.06 (0.04) | | 0.19 | 0.11 (0.05) | | 0.01 |
|  | APOE_Ɛ_4* Non-specific | 0.01 (0.04) | | 0.91 | -0.07 (0.05) | | 0.10 |
|  | Age | -0.04 (0.02) | | 0.006 | -0.02 (0.02) | | 0.19 |
|  | Sex | 0.24 (0.16) | | 0.14 | 0.18 (0.17) | | 0.29 |
|  | Years of education | -0.02 (0.02) | | 0.48 | 0.003 (0.02) | | 0.89 |

Note: DV, dependent variable; IV, independent variable; APOE _Ɛ_4, Apolipoprotein _Ɛ_4; FH, family history.

Supplementary Table 4c. CAIDE - Regression coefficient for Verbal and Visuospatial Functions, and Short-Term (conjunctive) Memory (Component 3) at Baseline & Follow-up

|  |  | Baseline | | | Follow-up | | |
| --- | --- | --- | --- | --- | --- | --- | --- |
|  | Model summary | R^2^ | F | p | R^2^ | F | p |
|  |  | 0.05 | 3.84 | 0.01 | 0.02 | 1.24 | 0.30 |
| DV | IV | β (SE) | | p | β (SE) | | p |
| Component 3 | CAIDE | -0.08 (0.03) | | 0.01 | -0.05 (0.03) | | 0.07 |
|  | Specific | -0.01 (0.02) | | 0.70 | -0.003 (0.02) | | 0.88 |
|  | Non-Specific | 0.02 (0.02) | | 0.21 | 0.01 (0.02) | | 0.70 |

DV, dependent variable; IV, independent variable; CAIDE, Cardiovascular Aging and Dementia risk score.

Supplementary Table 4d CAIDE - Regression coefficient for Verbal and Visuospatial Functions, and Short-Term (conjunctive) Memory (Component 3) at Baseline & Follow-up including interaction terms.

|  |  | Baseline | | | | | Follow-up | | | | |
| --- | --- | --- | --- | --- | --- | --- | --- | --- | --- | --- | --- |
|  | Model summary | R^2^ | | F | p | | R^2^ | | | F | p |
|  |  | 0.07 | | 3.11 | 0.01 | | 0.03 | | | 0.92 | 0.47 |
| DV | IV | β (SE) | | | p | | β (SE) | | | | p |
| Component 3 | CAIDE | -0.07 (0.03) | | | 0.01 | | -0.06 (0.03) | | | | 0.08 |
|  | Specific | -0.01 (0.02) | | | 0.54 | | -0.001 (0.02) | | | | 0.94 |
|  | Non-Specific | 0.02 (0.02) | | | 0.24 | | 0.01 (0.02) | | | | 0.63 |
|  | CAIDE*Specific | -0.01 (0.01) | | | 0.16 | | 0.04 (0.01) | | | | 0.58 |
|  | CAIDE*Non-specific | 0.01 (0.01) | | | 0.12 | | -0.01 (0.01) | | | | 0.40 |

DV, dependent variable; IV, independent variable; CAIDE, Cardiovascular Aging and Dementia risk score.
